## Supplementary Table 1 for "Timing of parental depression on risk of child depression and poor educational outcomes: a population based routine data cohort study from Born in Wales, UK"

### Supplementary Table 1: Depression read codes

| Read code | Read description |
|---|---|
| E112. | Single major depressive episode |
| E1120 | Single major depressive episode, unspecified |
| E1121 | Single major depressive episode, mild |
| E1122 | Single major depressive episode, moderate |
| E1123 | Single major depressive episode, severe, without psychosis |
| E1125 | Single major depressive episode, partial or unspec remission |
| E1126 | Single major depressive episode, in full remission |
| E112z | Single major depressive episode NOS |
| E113. | Recurrent major depressive episode |
| E1130 | Recurrent major depressive episodes, unspecified |
| E1131 | Recurrent major depressive episodes, mild |
| E1132 | Recurrent major depressive episodes, moderate |
| E1133 | Recurrent major depressive episodes, severe, no psychosis |
| E1135 | Recurrent major depressive episodes,partial/unspec remission |
| E1136 | Recurrent major depressive episodes, in full remission |
| E1137 | Recurrent depression |
| E113z | Recurrent major depressive episode NOS |
| E118. | Seasonal affective disorder |
| E135. | Agitated depression |
| E2003 | Anxiety with depression |
| E204. | Neurotic depression reactive type |
| E291. | Prolonged depressive reaction |
| E2B.. | Depressive disorder NEC |
| E2B0. | Postviral depression |
| E2B1. | Chronic depression |
| Eu32. | [X]Depressive episode |
| Eu320 | [X]Mild depressive episode |
| Eu321 | [X]Moderate depressive episode |
| Eu322 | [X]Severe depressive episode without psychotic symptoms |
| Eu324 | [X]Mild depression |
| Eu32y | [X]Other depressive episodes |
| Eu32z | [X]Depressive episode, unspecified |
| Eu33. | [X]Recurrent depressive disorder |
| Eu330 | [X]Recurrent depressive disorder, current episode mild |
| Eu331 | [X]Recurrent depressive disorder, current episode moderate |
| Eu332 | [X]Recurr depress disorder cur epi severe without psyc sympt |
| Eu334 | [X]Recurrent depressive disorder, currently in remission |
| Eu33y | [X]Other recurrent depressive disorders |
| Eu33z | [X]Recurrent depressive disorder, unspecified |
| Eu341 | [X]Dysthymia |
| Eu412 | [X]Mixed anxiety and depressive disorder |
