## Supplementary figures and images for "Timing of parental depression on risk of child depression and poor educational outcomes: a population based routine data cohort study from Born in Wales, UK"

### Supplementary Figure 1

Supplementary Figure 1: Flow Diagram of inclusion and exclusion within the study

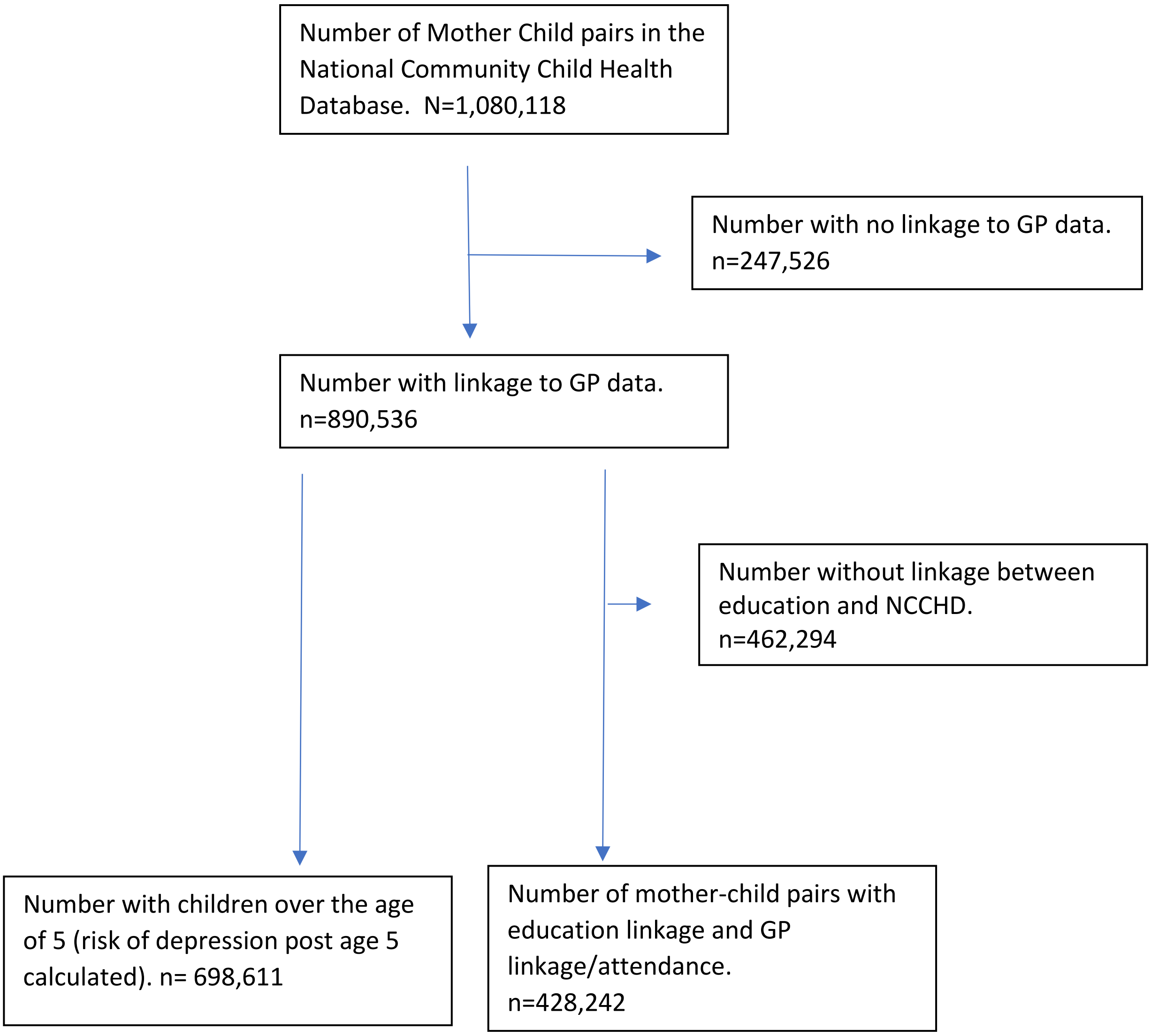
